# Variation in real world dementia risk profiles in 6171 adults drawn from the Australian CogDrisk website according to demographic characteristics

**DOI:** 10.1101/2025.07.24.25332109

**Authors:** Kaarin J. Anstey, Md Hamidul Huque, Ranmalee Eramudugolla, Meiwei Li, Marita Long, Scherazad Kootar, Michael Cartright, Claire MC O’Connor, Jeromey Temple

## Abstract

**Background:** We aimed to evaluate differences in dementia risk profiles according to age, birthplace and First Nations status.

**Methods:** The sample included 6171 (62% female, <1% non-binary/other) middle-aged and older adults who self-administered the CogDrisk assessment online and consented to their data being used for research from May 2023 to May 2024. The majority were aged 40-64 (‘midlife’; 67.9%) or aged 65-96 (‘later-life’; 25.8%); 6.3% were aged 18-40. More than a third (36.2%) were born overseas, 95% resided in Australia, and 66.0% had a university degree.

**Results:** Key midlife risk factors were hypertension 27.7%, pesticide exposure 24.6%, high cholesterol 25.1%, head injury 15.3%, diabetes 7.6%, and atrial fibrillation (afib) 7.3%. Key later-life risk factors were hypertension 45.4%, pesticide exposure 33.0%, afib 18.3% head injury 16.4%, and diabetes 11.7%. Midlife respondents from the Pacific and New Zealand (NZ) had high rates of vascular risk factors and those from NZ reported high rates of head injury and depression. The highest rates of heavy drinking and insufficient physical activity were reported by respondents from the UK and Asia respectively. Using published estimates from validation studies, 27% of those aged 65+ had a high dementia risk (CogDrisk score>12).

**Conclusions:** High rates of multiple risk factors were identified and varied according to age group and birthplace. This study provides evidence for tailoring risk reduction intervention and advice by age and birthplace.

## Introduction

Dementia is the second leading cause of death in Australia, and cost Australia $3.7 billion in 2020-2021[1]. Without any intervention, the rates and costs of dementia will increase astronomically due to population ageing. Over the past 15 years, there has been an accumulation of evidence on modifiable risk factors, indicating the potential for risk reduction to influence future rates of incident dementia. The Lancet commission [2] suggested that up to 45% of dementia cases world-wide could be attributable to modifiable risk factors for dementia, with individual country analyses showing similar if not higher estimates [3–5]. The risk factors for dementia are numerous and affect large numbers of people, across multiple influences on human health, including lifestyle, chronic disease, environmental exposures and genetics [6]. Patterns of risk also vary across demographic characteristics [7, 8] including racial and ethnic background. However, many studies aggregate data across demographic groups, leading to loss of information on the specific risk profiles for ethnically diverse and migrant groups who may have different lifestyle and medical conditions to the most widely represented cultural backgrounds in large studies. We aimed to evaluate dementia risk factor patterns according to ethnic and cultural background among a large sample of primarily Australian adults who completed a validated risk assessment tool [9, 10] in a naturalistic, cross-sectional study

### Methods

#### Sample

Data were drawn from the CogDrisk database hosted at Neuroscience Research Australia. Respondents (n = 6545) who consented for their data to be stored and used for research and completed the CogDrisk assessment between May 2023 to May 2024 were included. Only one occasion of measurement was included for each respondent. The collection of research data via the website for research was approved by the UNSW Human Research Ethics Committee. Data from 135 respondents were deleted due to missing data in age and 239 due to missing data in gender information. Of respondents who provided information on their address, 95% (n=2212) were from Australia, 49 (2.2%) from USA, 14 (0.5%) from UK, 11 from China (0.5%) and 10 from New Zealand. Among other countries we have data from Canada (5), Finland (1), Ireland (6), Malaysia (1), Mexico (1), Poland (1), Portugal (2), Romania (2), Spain (2), United Arab Emirates (1).

#### Measures

The CogDrisk is a 91-item questionnaire that includes validated scales and items to assess the presence of risk factors that have been reliably linked to dementia in cohort studies [9]. The term ‘risk factor’ in this study therefore refers to those factors assessed by the CogDrisk which are established as dementia risk factors in the wider literature. The average time required to complete the CogDrisk questionnaire was 15.3 minutes, while the CogDrisk Short Form (CogDrisk-SF) was completed in an average of 9.8 minutes[11]. It includes the following measures:

#### Demographic characteristics

Age in years, gender (Male, Female, Non-binary, Other identity, Prefer not to say), country of birth (Australia, England, China, New Zealand, India, Italy, Vietnam, Philippines, Other), language, employment status, highest education, and relationship status were recorded. For those born in Australia, identification as Aboriginal, Torres Strait Islander, both or neither was also recorded.

#### Psychological risk factors

The 10-item Centre for Epidemiological Studies Depression Scale (CES-D-10) [12] was used to screen for depression.

#### Behavioural risk factors

Smoking, drinking, fish consumption (diet), physical activity, and cognitive engagement were included to assess behavioural risk for dementia. Participants were asked whether they have ever smoked cigarettes, cigars, pipes or any other tobacco products (yes/no) or currently smoke cigarettes, cigars, pipes or any other tobacco products (yes/no). Participants were asked how many standard drinks they had on a typical day. For fish intake, participants were asked how often they eat fish or seafood that is not deep-fried (rarely, 1-3 times per month, once a week, 2-3 times per week, 4 or more times per week). The International Physical Activity Questionnaire short form (IPAQ-SF) [13] was used to assess physical activity. Participants were asked about time spent walking, in moderate and vigorous-intensity physical activity in the last 7 days. For cognitive engagement, participants were asked about activities they performed during leisure and work time (e.g., reading, play games, writing letters/emails etc).

#### Metabolic risk factors

Overweight was categorised as having a body mass index (BMI) ranging from 25.0 to under 30.0 kg/m², while obesity was defined as a BMI of 30.0 kg/m² or higher. These classifications were determined using self-reported height and weight, following the WHO guidelines (WHO, 2019). For diabetes and abnormal cholesterol, participants were asked if a doctor or other health professional had ever told them that they had the conditions (yes/no/don’t know).

#### Cardiovascular (CV) conditions

In the CogDrisk tool, participants were asked whether a doctor or other health professional had ever told them that they had any of the following CV conditions: high blood pressure, stroke or transient ischemic attack (TIA) (yes/no/don’t know).

#### Chronic health conditions

For chronic health conditions, participants were asked whether a doctor or other health professional had ever told them that they had any of the following conditions: head injury, atrial fibrillation, arrhythmias or hearing impairment (yes/no/don’t know).

#### Environmental risk factors

Pesticide exposure was included to assess environmental risk for dementia. Participants were asked whether they had ever been involved with mixing, applying or loading any pesticide, herbicide, weed killers, fumigants or fungicides (yes/no/don’t know).

### Dissemination activities related to dementia risk assessment using the CogDrisk website

A range of discrete dissemination strategies were used at the time of CogDrisk roll-out [14]. Information on CogDrisk was made available to the general public through media articles in 2023 and through podcasts for General Practitioners (GPs). To train and educate key stakeholders [15], information on CogDrisk was provided to GPs via professional development workshops in 2023. These included workshops/lectures and webinars conducted by Dementia Training Australia, Dementia Australia and by HealthEd which is the largest provider of professional development training for GPs in Australia. In 2023, more than 10,000 GPs participated in workshops, lectures or webinars at which the CogDrisk tool was described. More than 2,500 GPs also enrolled in a Continuing Professional Development activity that involved implementing the CogDrisk in their clinical practice for at least 5 patients. Finally, general enquiries about CogDrisk were possible via an email address on the website, monitored by the research team.

## Results

Demographic characteristics of the respondents (n=6171, 62% female, <1% non-binary/other) who consented to their data being saved and used for research is presented in Table 1. Among respondents, 6.3% were aged 18-39 years (early-life), 67.9% were aged 40-64 (midlife) and 25.8% were aged 65-96 (later-life). Less than 1% of the sample identified as Aboriginal, Torres Strait Islander or both. Approximately 35% were born overseas, which is relatively consistent with national estimates for Australia of approximately 30% [16]. Over three quarters of participants reported speaking only English at home, which again is relatively consistent with unadjusted 2021 Census of Population and Housing figures [17]. Of those not born in Australia, Italy was the most common country of birth, followed by UK, India, China, and New Zealand. University level education was reported by 66.0% which is higher than the Australian average reported in 2022 of 32% [18]. Supplementary Table S1 shows the demographic characteristics according to gender identification. Figure 1 shows the distribution of respondents geographically and illustrates the national reach of the implementation process.

**Figure 1.**
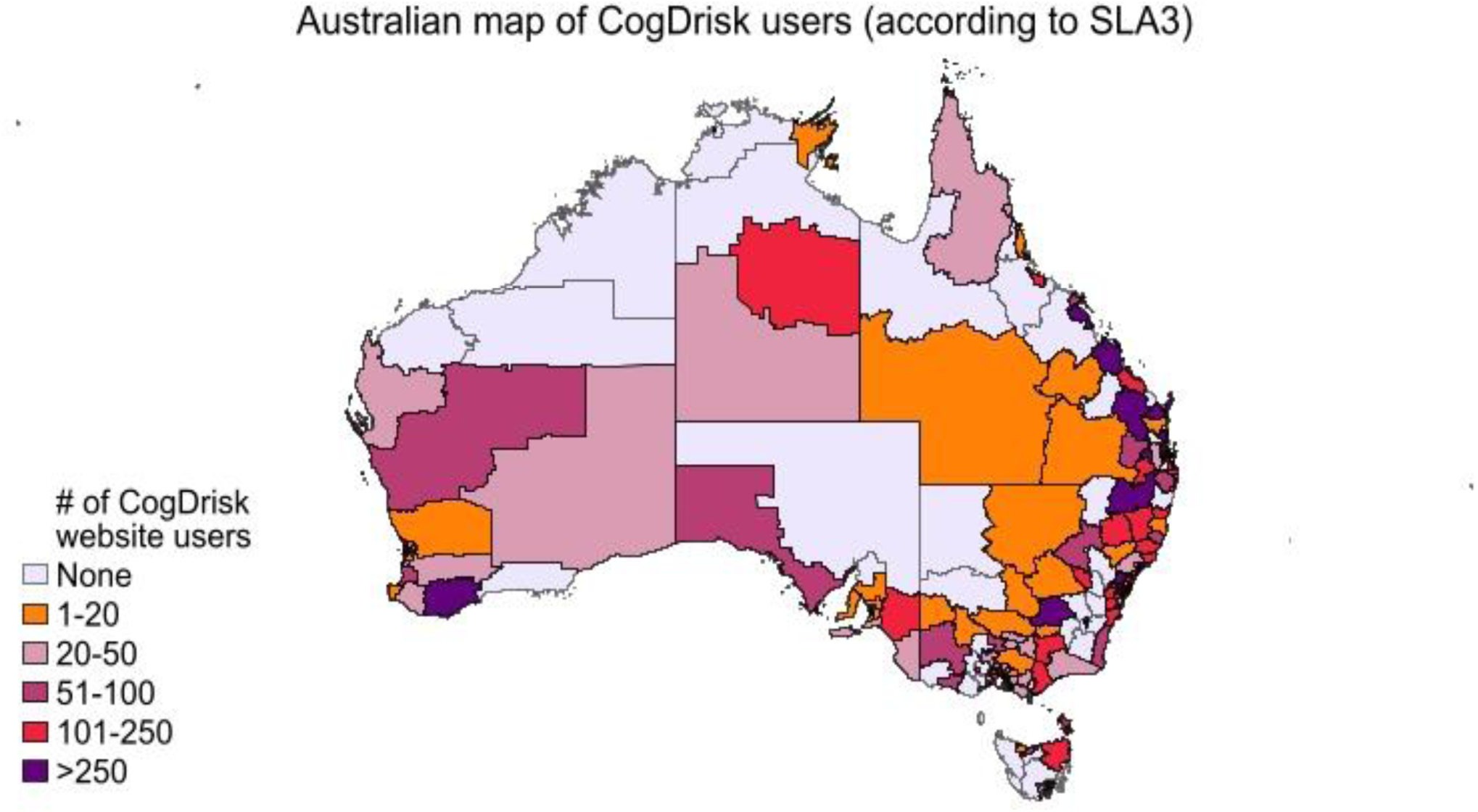
Map of distribution of CogDrisk users across Australia

**Table 1:**
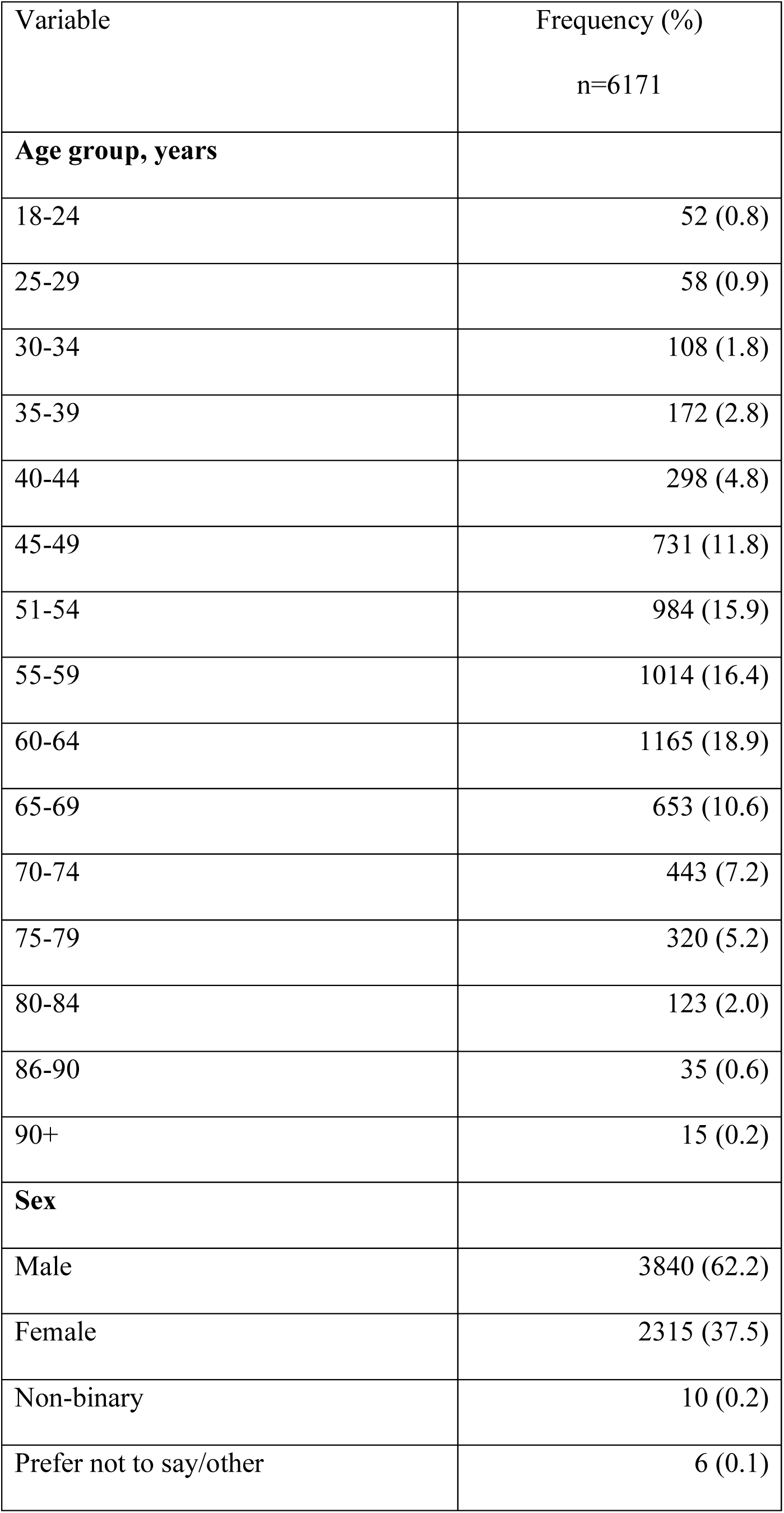
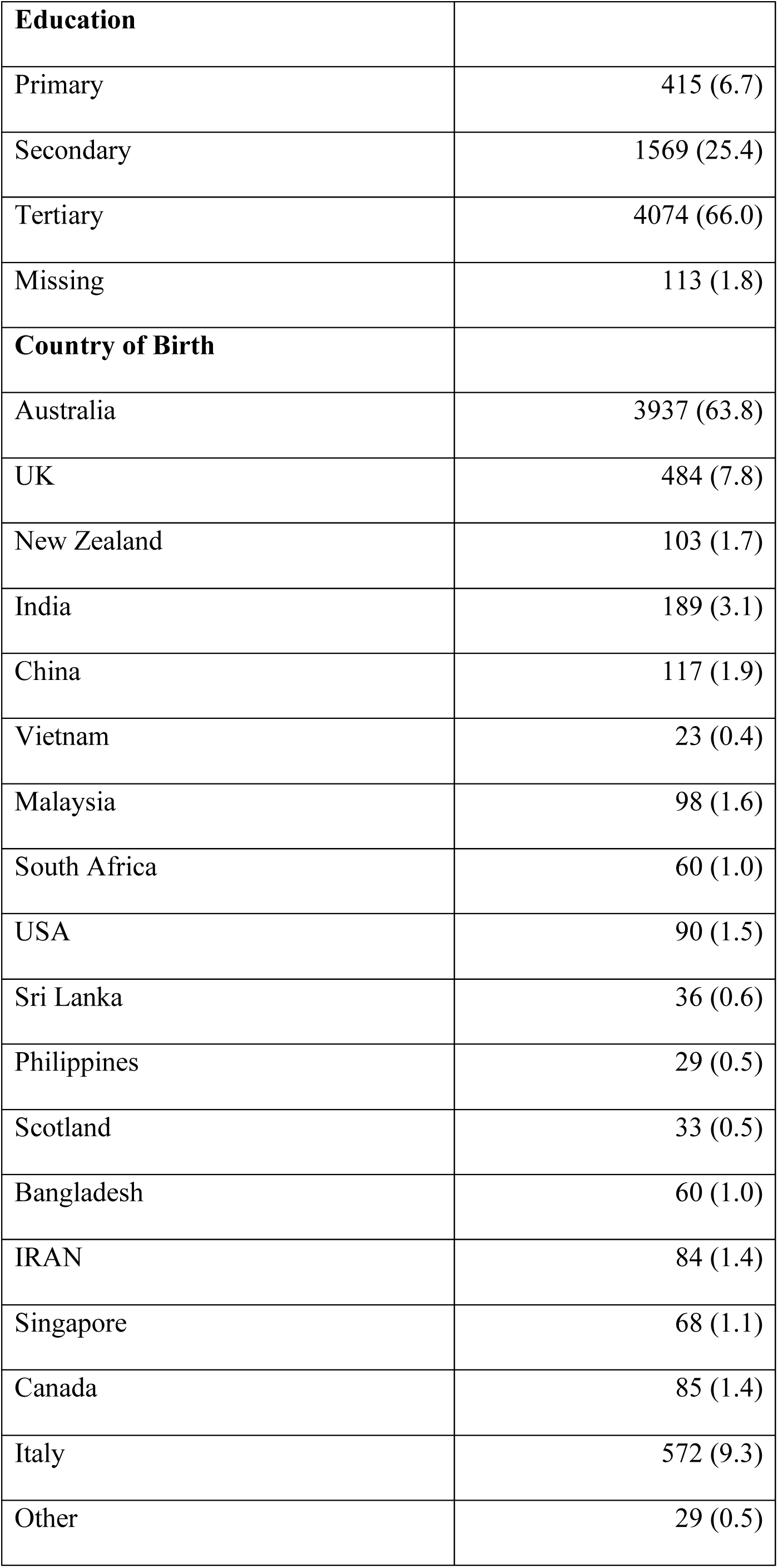
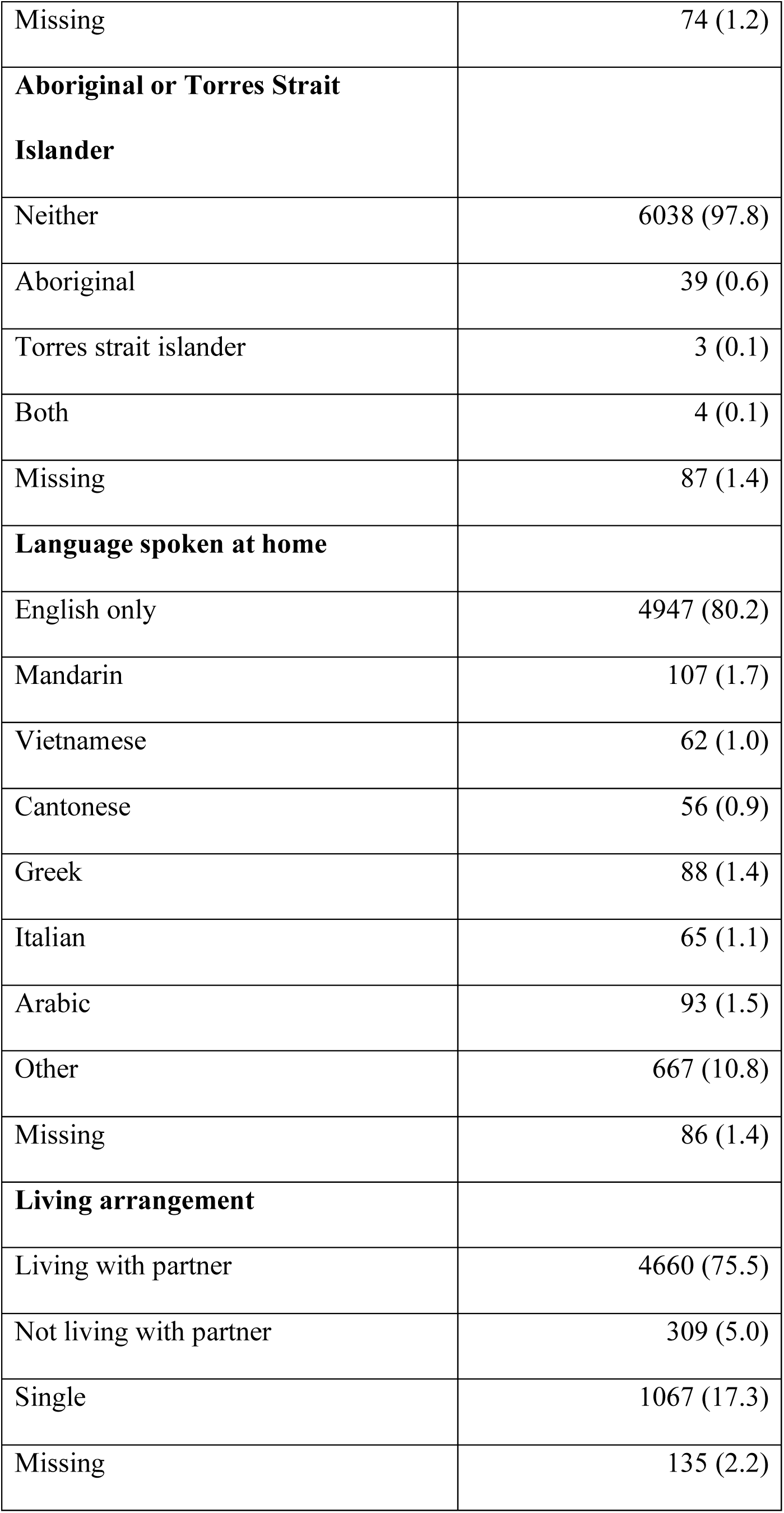
Descriptive characteristics of the users of CogDrisk online tool.

| Variable | Frequency (%)<br>n=6171 |
| --- | --- |
| **Age group, years** |  |
| 18-24 | 52 (0.8) |
| 25-29 | 58 (0.9) |
| 30-34 | 108 (1.8) |
| 35-39 | 172 (2.8) |
| 40-44 | 298 (4.8) |
| 45-49 | 731 (11.8) |
| 51-54 | 984 (15.9) |
| 55-59 | 1014 (16.4) |
| 60-64 | 1165 (18.9) |
| 65-69 | 653 (10.6) |
| 70-74 | 443 (7.2) |
| 75-79 | 320 (5.2) |
| 80-84 | 123 (2.0) |
| 86-90 | 35 (0.6) |
| 90+ | 15 (0.2) |
| **Sex** |  |
| Male | 3840 (62.2) |
| Female | 2315 (37.5) |
| Non-binary | 10 (0.2) |
| Prefer not to say/other | 6 (0.1) |
| **Education** |  |
| Primary | 415 (6.7) |
| Secondary | 1569 (25.4) |
| Tertiary | 4074 (66.0) |
| Missing | 113 (1.8) |
| **Country of Birth** |  |
| Australia | 3937 (63.8) |
| UK | 484 (7.8) |
| New Zealand | 103 (1.7) |
| India | 189 (3.1) |
| China | 117 (1.9) |
| Vietnam | 23 (0.4) |
| Malaysia | 98 (1.6) |
| South Africa | 60 (1.0) |
| USA | 90 (1.5) |
| Sri Lanka | 36 (0.6) |
| Philippines | 29 (0.5) |
| Scotland | 33 (0.5) |
| Bangladesh | 60 (1.0) |
| IRAN | 84 (1.4) |
| Singapore | 68 (1.1) |
| Canada | 85 (1.4) |
| Italy | 572 (9.3) |
| Other | 29 (0.5) |
| Missing | 74 (1.2) |
| **Aboriginal or Torres Strait Islander** |  |
| Neither | 6038 (97.8) |
| Aboriginal | 39 (0.6) |
| Torres strait islander | 3 (0.1) |
| Both | 4 (0.1) |
| Missing | 87 (1.4) |
| **Language spoken at home** |  |
| English only | 4947 (80.2) |
| Mandarin | 107 (1.7) |
| Vietnamese | 62 (1.0) |
| Cantonese | 56 (0.9) |
| Greek | 88 (1.4) |
| Italian | 65 (1.1) |
| Arabic | 93 (1.5) |
| Other | 667 (10.8) |
| Missing | 86 (1.4) |
| **Living arrangement** |  |
| Living with partner | 4660 (75.5) |
| Not living with partner | 309 (5.0) |
| Single | 1067 (17.3) |
| Missing | 135 (2.2) |

### Overall risk, and risk factors by age group, gender and education

The mean (standard deviation, SD) CogDrisk score was 2.8 (4.3) for midlife, 7.7 (5.8) in later life. Among later-life participants, this figure is 8.1 (5.9) and 7.2 (5.6) for male, and female participants, respectively. Using estimates from published validation studies, 19.4% of the sample aged 65 and above had high dementia risk (CogDrisk score>12). Figure 2 shows the frequencies of modifiable risk factors by age and sex. This shows a normal distribution of the number of risk factors, with the mode being approximately 4-5 risk factors (excluding age, and sex).

**Figure 2.**
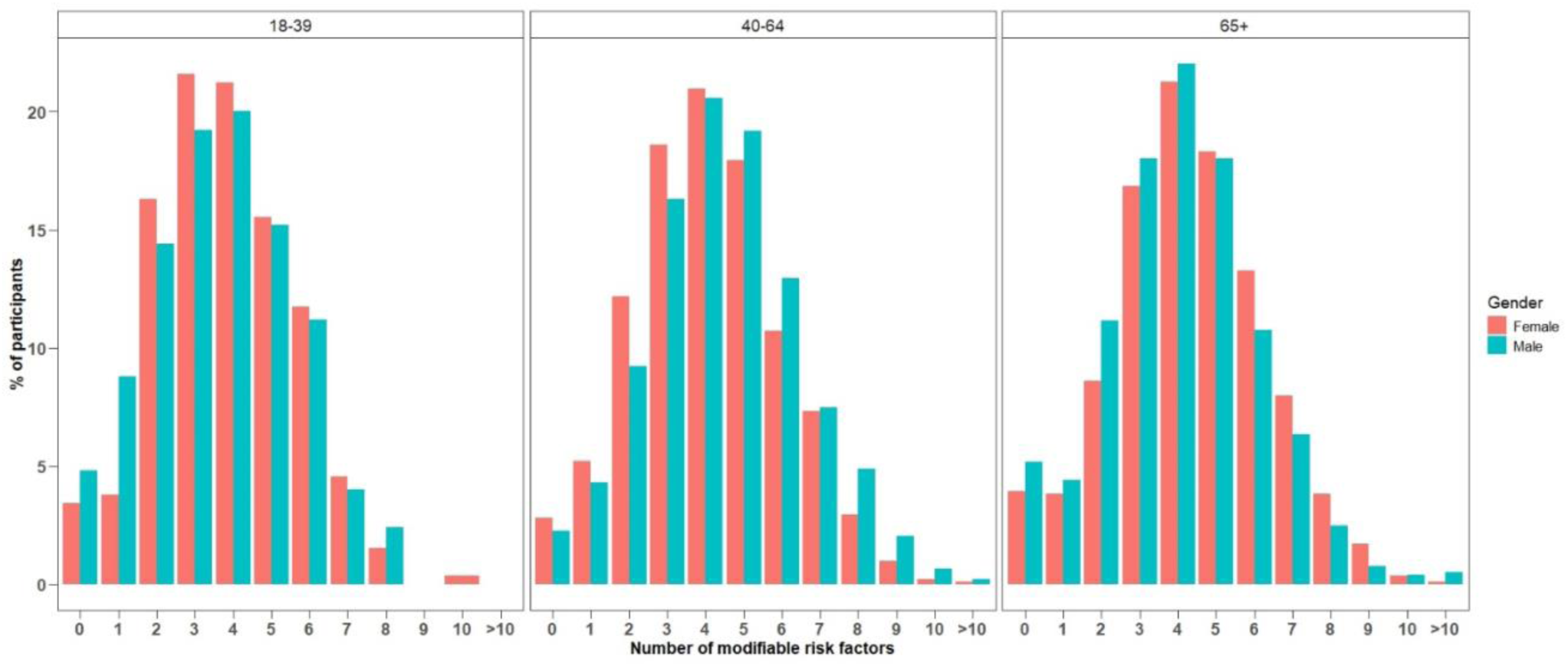
Number of modifiable factors in each age group by sex

Table 2 reports the frequency of respondents reporting each modifiable dementia risk factor by three stages of life (early life: age 18-39 years, midlife: age 40-64 years and later-life: age 65 and older). Across the whole sample of midlife adults, risk factors were relatively high with the main factors being: hypertension (27.7%), high cholesterol (25.1%), pesticide exposure (24.6%), head injury (15.3%), diabetes (7.6%), atrial fibrillation (afib) (7.3%) and current smoking (5.4%). In later-life, key risk factors were hypertension (45.4%), pesticide exposure (33.0%), high cholesterol (23.8), afib (18.3%), head injury (16.4%), and diabetes (11.7%).

**Table 2:** Risk factors according to three age stages of adulthood.

|  | Early Adulthood<br>(18-39 years)<br>n=390 (%) | Midlife<br>(40-64 years)<br>n=4192 (%) | Late life<br>(65 years and above)<br>n=1589 (%) |
| --- | --- | --- | --- |
| **Obesity** |  |  |  |
| Underweight | 12 (3.1) | 43 (1.0) | 18 (1.1) |
| Normal | 201 (51.5) | 1513 (36.1) | 579 (36.4) |
| Overweight | 85 (21.8) | 1432 (34.2) | 558 (35.1) |
| Obese | 79 (20.3) | 1056 (25.2) | 304 (19.1) |
| Missing | 13 (3.3) | 148 (3.5) | 130 (8.2) |
| **High Cholesterol** | 39 (10.0) | 1051 (25.1) | 378 (23.8) |
| **Diabetes** | 6 (1.5) | 319 (7.6) | 186 (11.7) |
| **Stroke** | 1 (0.3) | 78 (1.9) | 72 (4.5) |
| **Head Injury** | 59 (15.1) | 642 (15.3) | 261 (16.4) |
| **Hypertension** | 36 (9.2) | 1160 (27.7) | 722 (45.4) |
| **Blood Pressure medication** | 8 (2.1) | 859 (20.5) | 645 (40.6) |
| **Atrial Fibrillation** | 20 (5.1) | 304 (7.3) | 290 (18.3) |
| **Insomnia** | 148 (37.9) | 1381 (32.9) | 378 (23.8) |
| **Depression** | 189 (48.5) | 1256 (30.0) | 309 (19.4) |
| **Low Physical activity** | 42 (10.8) | 379 (9.0) | 132 (8.3) |
| **Cognitive engagement** |  |  |  |
| Low | 147 (37.7) | 1331 (31.8) | 354 (22.3) |
| Middle | 191 (49.0) | 2226 (53.1) | 894 (56.3) |
| High | 14 (3.6) | 389 (9.3) | 203 (12.8) |
| missing | 37 (9.5) | 246 (5.9) | 138 (8.7) |
| **Loneliness** | 229 (58.7) | 3007 (71.7) | 1168 (73.5) |
| **Fish intake (serve/wk)** |  |  |  |
| 0 | 100 (25.6) | 708 (16.9) | 176 (11.1) |
| 0.5 | 108 (27.7) | 1051 (25.1) | 315 (19.8) |
| 1 | 89 (22.8) | 1241 (29.6) | 496 (31.2) |
| 2.5 | 35 (9.0) | 694 (16.6) | 355 (22.3) |
| 4 | 8 (2.1) | 155 (3.7) | 54 (3.4) |
| Missing | 50 (12.8) | 343 (8.2) | 193 (12.1) |
| **Smoking status** |  |  |  |
| Non-smoker | 228 (58.5) | 2368 (56.5) | 796 (50.1) |
| Former smoker | 78 (20.0) | 1233 (29.4) | 571 (35.9) |
| Current smoker | 29 (7.4) | 227 (5.4) | 23 (1.4) |
| Missing | 55 (14.1) | 364 (8.7) | 199 (12.5) |
| **Alcohol drink** |  |  |  |
| Abstainer | 66 (16.9) | 739 (17.6) | 250 (15.7) |
| Moderate | 235 (60.3) | 2540 (60.6) | 835 (52.5) |
| Heavy drinker | 35 (9.0) | 528 (12.6) | 301 (18.9) |
| Missing | 54 (13.8) | 385 (9.2) | 203 (12.8) |
| **Pesticides exposure** | 50 (12.8) | 1031 (24.6) | 525 (33.0) |

Figure 3 and supplementary Table S2 report the frequency of respondents reporting each modifiable dementia risk factor by age and gender. Striking age differences were seen for depression, which was far higher among the youngest age group, and within that group, higher for females than males. Of note is the high rate of loneliness in all age groups, with 58.7% in the younger adults, 71.7% in the middle aged and 73.5% in the older adults. Notably there were sex differences in prevalence of various risk factors across different age-groups. A significantly higher proportion of males were overweight, and reported being lonely compared with females across all age-groups. Among the participants from midlife and late-life, a significantly higher proportion of males compared to females reported diabetes, hypertension, atrial fibrillation, smoking, and heavy drinking (Supplementary Table S2).

**Figure 3:**
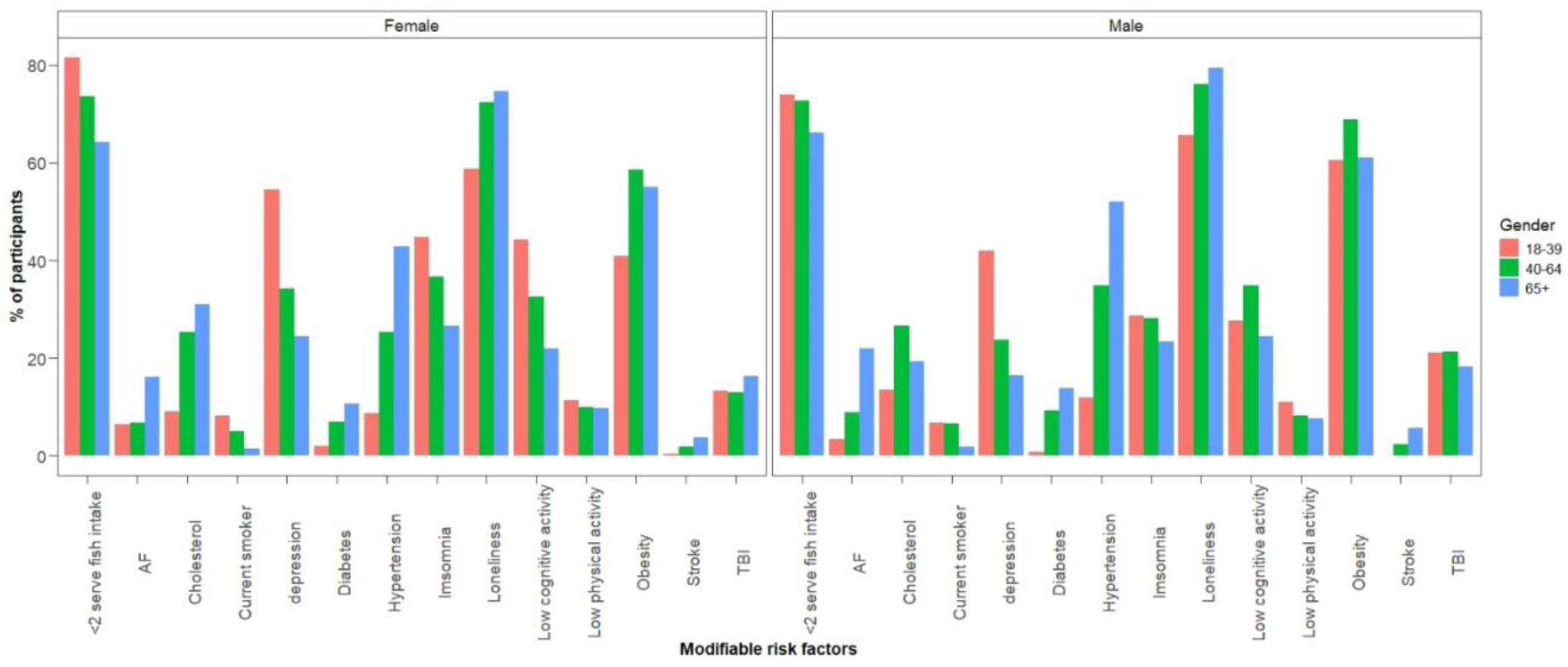
Frequency distribution of modifiable risk factors by age-group and gender TBI, Traumatic brain injury

Overall, the rates of smoking and stroke in the sample were low. Dementia risk was higher among respondents with lower levels of education. Removing education from the scoring of CogDrisk (to avoid confounding), showed that 39.5% of those with secondary education, 27.9% with higher secondary or equivalent and 18.5% of those tertiary education had high risk of dementia respectively, using previously published cutoffs.

### Risk factors by country or region of origin

There was notable variation among the rates of risk factors according to country or region of birth and First Nations Australians although samples sizes were very small for some groups (Tables 3, 4, Supplementary Table S3). Among the midlife respondents, those born in the Pacific region had the highest rates of obesity, diabetes, hypertension, and smoking, while those from East Asia reported the highest rates of high cholesterol. Midlife respondents from New Zealand also had high rates of obesity, head injury, hypertension, afib, and depression. Respondents from the UK had the highest rates of heavy drinking. Respondents from southeast Asian and other Asian regions reported the highest rates of insufficient physical activity.

**Table 3:**
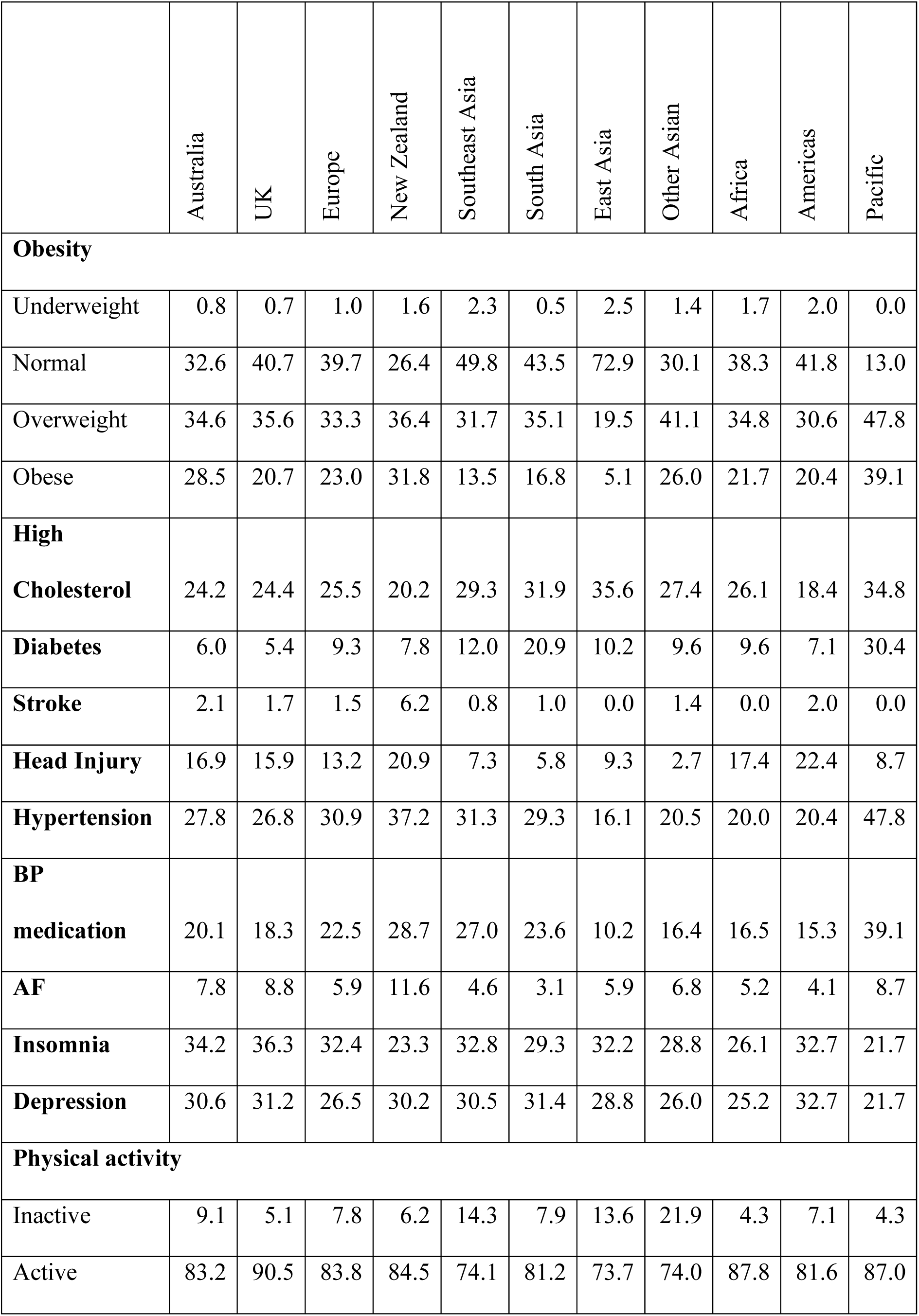
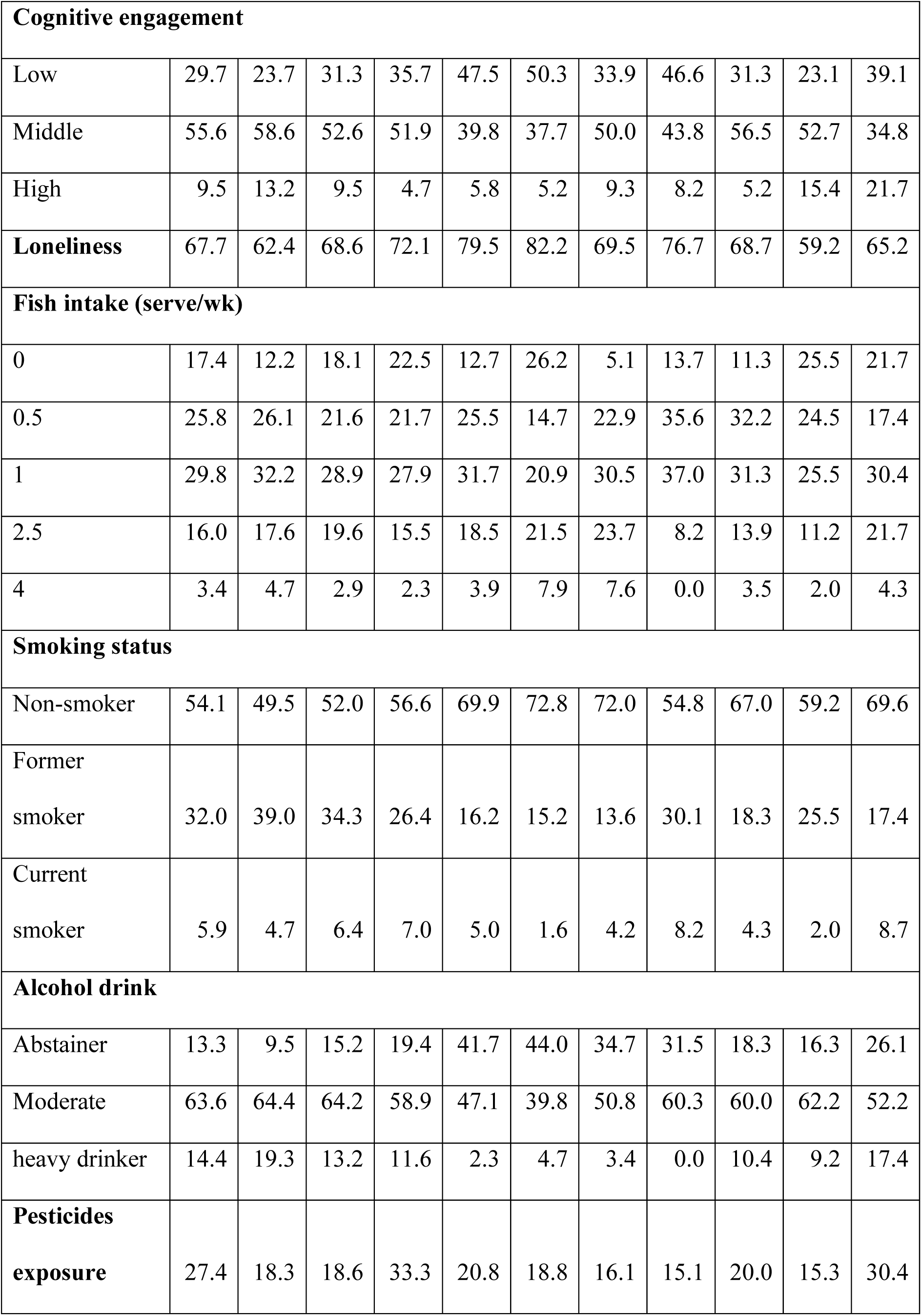
Dementia Risk/Protective Factors by Country of Birth in Participants Aged 40-64.

**Table 4:**
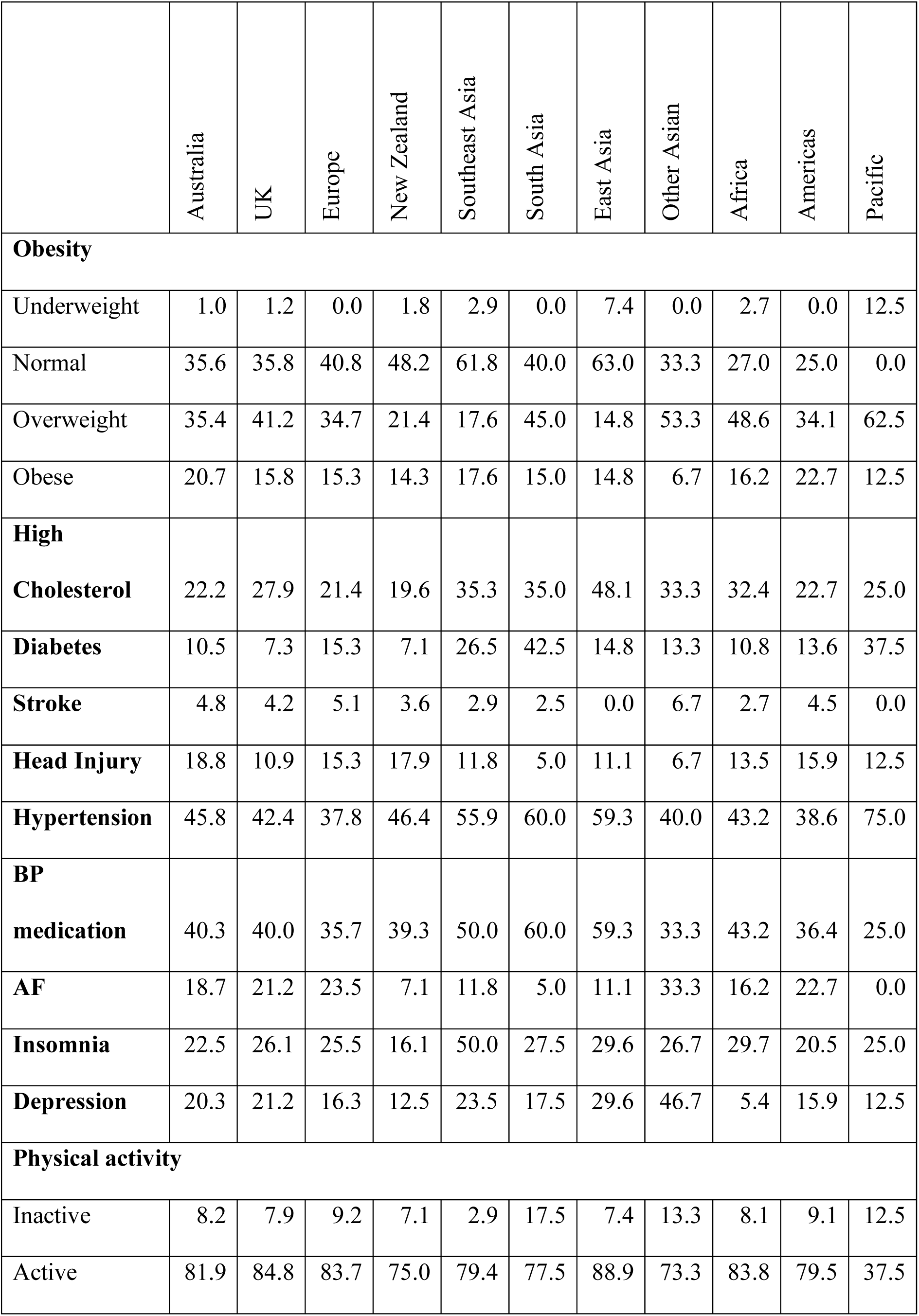
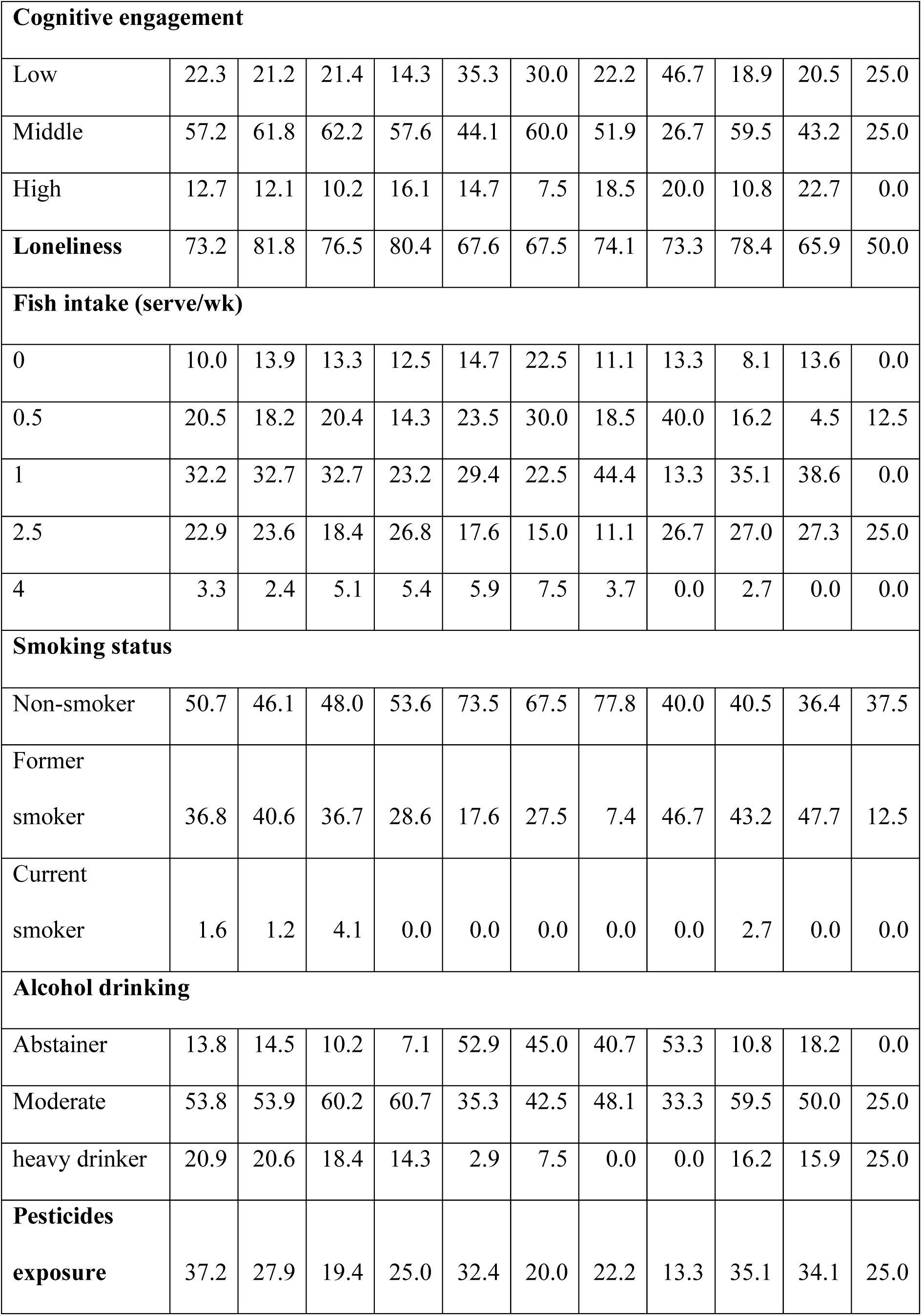
Dementia Risk/Protective Factors by Country of Birth in Participants Aged 65+.

Among the later-life respondents, obesity rates were generally lower than among the midlife respondents and the highest rate was observed in respondents from the Americas. Hypertension was highest among respondents from Asia, and depression was notably high among the ‘other Asian’ group. South Asian respondents reported the highest rates of low physical activity and low cognitive activity. Smoking rates were highest among later life adults from the UK, Africa, and Other Asian regions.

Overall, among midlife and late-life adults, those born overseas had higher proportions of high cholesterol and diabetes compared with the Australian born respondents. On the other hand, Australian born participants reported higher rates of head injury and exposure to pesticides compared with their counterparts (Supplementary Table S3).

First Nations Australian respondents reported higher rates of vascular risk factors (obesity, high cholesterol, diabetes, hypertension), and higher rates of depression and low cognitive engagement compared with non-First Nations Australians (Supplementary Table S4).

## Discussion

To our knowledge this is the first real world data following broad implementation of a dementia risk assessment tool. Our study identified significant variation in the types of risk factors reported in midlife and later life, and by country of birth. It must be recognised that this is not an epidemiological sample and as such is not representative of the population. The sample reflects adults who were referred by their GP or who self-referred to assess their own risk factors. However, consistent with representative national studies of Australians, we found that a higher proportion of males compared with females were overweight and obese [19], and have diabetes, and hypertension[20]. Therefore, the real-world implementation of this dementia risk assessment tool will provide valuable data as the sample size grows over time. Considering this, the trends shown in the data provide useful information for policy makers and practitioners, indicating that specific communities may require more tailored risk reduction strategies or healthy lifestyle support. For example, middle aged adults from the Pacific reported high rates of vascular risk factors, many of which require medical management. Some Asian groups reported low physical and cognitive activity which indicates a need for lifestyle modification.

These differences in risk factors among birthplace groups are important given the significant demographic change in Australia’s older population. Recent modelling indicates a rapid evolution from an older population with a predominately European birthplace and background, to an increasing Asian born background [21]. This differential ageing and population growth results in considerable heterogeneity in future older populations living with dementia. Again, by mid-century, projections indicate a slight decrease in populations living with dementia among those born in Southern & Eastern Europe to over 600% increases amongst the South-East Asia, Southern & Central Asia, and Sub-Saharan Africa-born populations in Australia [22]. Similarly, within the Aboriginal and Torres Strait Islander population, although this population has a younger age structure than the non-Indigenous population, the speed of numerical ageing is significant – over 800% among those aged over 85 by mid-century [23]. This demographic change translates into a 4.5 – 5.5 fold increase in the number of Aboriginal people living with dementia by mid-century [24]. How differential risk factors are addressed in high population ageing groups among different cultural groups requires considerable policy attention.

One particularly concerning result from our study is the high rate of depression reported by all groups. In the CogDrisk, depression is measured using the Centre for Epidemiological Studies Depression Scale and clinically significant depression is identified using the clinical cutoff of 20. Hence these results are valid and are higher than expected when compared with national statistics according to which 6.2% of males and 7.1% of females aged 45-54 reported experiencing an affective disorder [25]. It is possible that this reflects selection bias in the sample, with those experiencing higher rates of depression being more likely to undertake the assessment or more likely to be identified by GPs as at risk and that the CES-D captures more cases than the Australian Bureau of Statistics (ABS) methodology.

Another important finding was the high rates of adults completing the questionnaire who were classified as high risk for dementia according to studies that validated CogDrisk [26]. This indicates that the implementation of this risk assessment tool is leading to identification of adults with potential to benefit from dementia risk reduction interventions. In addition, the respondents in this study had high rates of modifiable risk factors, indicating a large opportunity for risk reduction interventions. Participants were provided with educational materials tailored to their dementia risk score once they completed the assessment. An important next step in this research will be to encourage people to re-assess their dementia risk after a period of time with the hope that people may feel empowered to address their identified dementia risk factors [27]. The discrete implementation strategies used in this study were an important initial step to drive uptake of CogDrisk. Specifically, GPs are often recognised to be gatekeepers for health initiatives [28], therefore, they were recognised as an important stakeholder group to target in this study [14]. However, it has been recognised that engaging GPs in new initiatives or research can be challenging [29]. Therefore, leveraging educational meetings for GPs via established national educational organisations to disseminate information about the CogDrisk was a useful implementation strategy to help with reach and legitimisation of the new tool [15].

Limitations to the study include lack of information on whether individuals were referred by their GP or were self-referred and the cross-sectional design. Data were only available from those consenting to research so the results may not reflect the risk profiles of the full sample who completed the risk score. Individuals with more risk factors may be more likely to seek out the tool or be referred by their GP. The sample had a higher level of university education than the Australian average, and this may reflect bias in university educated adults being more aware of research studies and hence more prepared to participate in them. Moreover, our measure of cultural diversity was limited to country of birth in addition to identification as an Aboriginal and Torres Strait Islander. Additional studies are sorely needed with more detailed measures of ethnicity to examine differential risk factors and association with dementia risk. For example, for the New Zealand born participants in our study, we were unable to measure those who identify as Maori. This distinction is important, dementia etiology may be influenced by ethnicity [30]. The numbers of respondents within some groups were small and results to not necessarily reflect the broader levels of risk within population groups as sampling was not representative. A repeated measures design would also provide valuable information on the stability of observed risk profiles.

We conclude that there is enormous scope to reduce future dementia risk in the current Australian population. However, risk reduction strategies need to be tailored to age, gender, cultural and ethnic background, and culturally tailored and appropriate strategies are needed for First Nations Australians.

## Data Availability

All data produced in the present study are available upon reasonable request to the authors

## Abbreviations

*afib*: Atrial fibrillation
*NZ*: New Zealand
*CogDrisk-SF*: CogDrisk Short Form
*CES-D-10*: 10-item Centre for Epidemiological Studies Depression Scale
*IPAQ-SF*: International Physical Activity Questionnaire short form
*BMI*: Body mass index
*CV*: Cardiovascular
*TIA*: Transient ischemic attack
*GP*: General Practitioner
*SD*: Standard deviation
*ABS*: Australian Bureau of Statistics

## Declarations

### Ethics approval and consent to participate

Ethics approval is provided by the University of New South Wales Human Research Ethics Committee (UNSW HREC; protocol number HC200108). All data are de-identified and stored on a secure server at Neuroscience Research Australia. All the participants provided informed consent to the original data collection.

### Consent for publication

Not applicable.

### Availability of data and materials

The study data can be accessed by contacting the corresponding author.

### Competing interests

The authors declare no competing interests.

### Funding

The development of the CogDrisk website was funded by a NeuRA Discovery Grant. Anstey and Huque are funded by FL190100011. O’Connor is funded by a Dementia Centre for Research Collaboration Postdoctoral Fellowship.

### Authors’ contributions

KJA obtained funding, conceived the project, provided supervision and drafted the manuscript. HH conducted the statistical analysis. SK and MC developed the website with input from KA, RE, HH, and ML. CO provided expertise in implementation, ML provided clinical expertise and JT provided expertise on demographic trends. All authors provided critical input into the manuscript.

## Acknowledgements

Programming support was provided by NeuRA IT.

## Notes

### Competing Interest Statement

The authors have declared no competing interest.

### Author Declarations

Ethics committee of University of New South Wales gave ethical approval for this work

